# Structural Cardiac Abnormalities, Ventricular Dysfunction Phenotypes, and Heart Failure Risk among Antiretroviral Therapy-treated People Living with HIV in South Africa

**DOI:** 10.64898/2026.06.04.26354960

**Authors:** Zaayid Omar, Abdullahi M. Ahmed, Julian Wolfson, Zuofu Huang, Msimelelo Mgidlana, Ashley Black, Marya Abd El Hadi, Olukayode O. Aremu, Tess Peterson, Ntobeko A. B. Ntusi, Graeme Meintjes, Mpiko Ntsekhe, Jason V. Baker, the PHIZA Study Team

**Author notes:** **Corresponding author** (name, email, complete address): Zaayid Omar, CIDRI-Africa Room 2.1 Wernher & Beit North Building, Institute of Infectious Disease & Molecular Medicine, University of Cape Town Faculty of Health Sciences, Anzio Rd, Observatory, Cape Town, South Africa, 7935. contributed equally to the development of this work.

## Abstract

**Background:** The manifestations of cardiovascular disease (CVD) among people with HIV (PWH) differ by region globally. While HIV disease is associated with increased atherosclerotic CVD risk in the global North, non-ischemic heart failure (HF) is more common in sub-Saharan Africa, the global HIV epicenter. We estimated the effect of treated HIV on the frequency and phenotype of HF and its cardiac precursors in South Africa (SA).

**Methods:** In an observational study, we recruited PWH on antiretroviral therapy (ART), age ≥40 years and people without HIV (PWoH) with similar distributions of age, sex, ethnicity, and hypertension, from a community clinic in Khayelitsha (Cape Town, SA). Procedures included a clinical assessment, echocardiography (Echo), and b-type natriuretic peptide (BNP) measure. Echo parameters defined structural abnormalities, left ventricle (LV) filling pressure, and LV systolic and diastolic dysfunction (DD). HF was defined by symptoms and/or BNP ≥35pg/mL and LV dysfunction, subcategorized as reduced, mildly reduced, or preserved ejection fraction (HFrEF, HFmrEF, and HFpEF). Comparisons by HIV status were adjusted for age, sex, hypertension, smoking, obesity, diabetes, elevated LDL-cholesterol, and hazardous alcohol use.

**Results:** Between September 2022 and August 2025, we enrolled 1008 PWH and 500 controls [median (Q1-Q3) age 48 years (43-53), 77% female]. Among PWH and controls respectively, 37% and 39% had hypertension, 21% and 25% were current smokers, 40% and 45% were obese, and 9% and 17% had diabetes. LV systolic dysfunction (1%) and HFrEF (1%) were rare, and undiagnosed HFpEF (8%) was the predominant HF phenotype. Compared to controls, PWH had higher odds of elevated LV mass index (LVMI) (OR 2.1; 95%CI 1.5-3.0) and DD (OR 1.4; 95%CI 1.0-2.0). Risk for elevated LVMI and DD was greatest among women with HIV, who also had an increased risk for undiagnosed HFpEF (OR 1.9; 95%CI 1.2-3.2), compared to women without HIV; effects which were not seen among men (p=0.051 for HIV*Sex interaction).

**Conclusions:** In a peri-urban SA community with a high burden of cardiometabolic risk factors, the frequency of abnormal structural and functional cardiac precursors of HFpEF was greater amongst ART-treated PWH. This was most pronounced amongst women with HIV, who also had increased risk of undiagnosed HFpEF.

**Clinical Perspective:** 1) What is new? *(maximum 100 words, formatted as 2-3 bullets)*

- In this community with a high prevalence of HIV, tuberculosis, and cardiometabolic risk factors, heart failure (HF) frequency (10%) was high and dominated by preserved ejection fraction (HFpEF) (8%).
- Echocardiographic defined structural and functional precursors of HFpEF were comparatively more frequent in people with HIV,
- Women with HIV were at highest risk of HFpEF.

2) What are the clinical implications? (maximum 100 words, formatted as 2-3 bullets).

- There is a need for resource efficient tools for screening and early detection of HFpEF in primary care clinics serving similar communities.
- The relationship between HIV, sex, and adiposity in HFpEF pathogenesis needs further study.

## Introduction

HIV remains a major contributor to morbidity and mortality in sub-Saharan Africa, where the majority of people with HIV (PWH) reside globally^1^. Widespread access to anti-retroviral therapy (ART) has led to prolonged survival and reduced risk for AIDS complications, but the relative proportion of deaths and disability attributable to age-related chronic conditions such as cardiovascular disease (CVD) has increased, and these conditions present at earlier ages.^2^ It has been estimated that ART-treated PWH have a twofold higher risk of developing both atherosclerotic CVD (ASCVD) events and heart failure (HF).^3^ However, epidemiologic data describing the changing spectrum of HIV-associated comorbid conditions largely arise from high income countries (HICs) in the global North, and the generalizability of these data to sub–Saharan Africa (SSA) is unclear.^4^

In SSA, data suggests that the spectrum and burden of CVD amongst both PWH and people without HIV (PWoH) is different from HICs, with non-ischemic CVD manifestations playing a more prominent role.^5–8^ Much of the published literature on HIV-related CVD from SSA predates current clinical practice with effective ART treatment, is based on descriptions of risk factor prevalence^9,10^, and/or relies on epidemiological models informed by data from HICs to estimate HIV-associated CVD risk in low-middle income countries.^3^ More recent data suggests that, when compared to HICs, SSA has much lower rates of ASCVD and its complications^5,11^ and higher rates of HF due to non-ischemic causes such as hypertension, primary valve pathology, and heart muscle disease.^12^ Importantly, when considering HIV-associated CVD between HICs and SSA, there are major differences in population characteristics, social determinants of health, and burden of non-traditional CVD risk factors such as tuberculosis.^13^

Here we report the primary findings from the Prevalence and Phenotype of HIV-HFpEF in South Africa (PHIZA) study. This study provides contemporary data within a front-line clinical setting in SSA on the contribution of HIV to HF risk, including the frequency of cardiometabolic risk factors influencing HF risk, the spectrum of structural and functional cardiac abnormalities, the frequency of undiagnosed clinical HF and the underlying phenotypes within a high HIV prevalence region in the global South.

## Methods

### Study Design and Population

PHIZA was an observational study of PWH and PWoH designed to determine the prevalence of major heart muscle and cardiac chamber abnormalities, and HF phenotypes in a SSA community with a high prevalence of HIV and tuberculosis. Study participants were enrolled at the Site B Community Health Center clinic in Khayelitsha, a low socio-economic peri-urban town outside of Cape Town, SA. Eligibility criteria included PWH who were 40 years and older, on ART for at least one year without interruption, and virally suppressed (HIV viral load level <200 copies/mL) on their most recent clinical laboratory evaluation. Control participants were PWoH from the same community with recruitment targeting similar frequencies of age ≥50 years, female sex, and hypertension status, to prevent large differences between groups in these potential confounders. Key exclusions included women who were pregnant or lactating, active tuberculosis or bacterial infection, and chemotherapy treatment for cancer within the prior six months.

The study was approved by the health research ethics and human subject research committees at the University of Cape Town [HREC Reference 119/2022] and Hennepin Healthcare Research Institute, respectively. All participants underwent verbal and written informed consent procedures in isiXhosa, Afrikaans or English, as appropriate.

### Study Assessments

Participants underwent clinical, cardiac, and laboratory assessments as described in **Figure 1**. Symptoms suggestive of HF were solicited via a structured 5-question checklist that asked about recent shortness of breath, dyspnea on exertion, nocturnal dyspnea, fatigue, and lower limb swelling (see **Supplemental Material** for full list of Supplemental Methods). Additional cardiac assessments were conducted in the clinic setting and included a blood pressure, electrocardiogram, echocardiogram (ECHO), and point of care blood testing for B-type natriuretic peptide (BNP) and Troponin-I levels (Quidel Alere™ Triage® MeterPro). ECHO imaging was performed by an experienced cardiac ultrasound technician and approximately one-third of images (n=425) were reviewed remotely by a cardiologist for quality assurance, including all participants with features of possible diastolic dysfunction and a random sample of patients without LV dysfunction.

**Figure 1:**
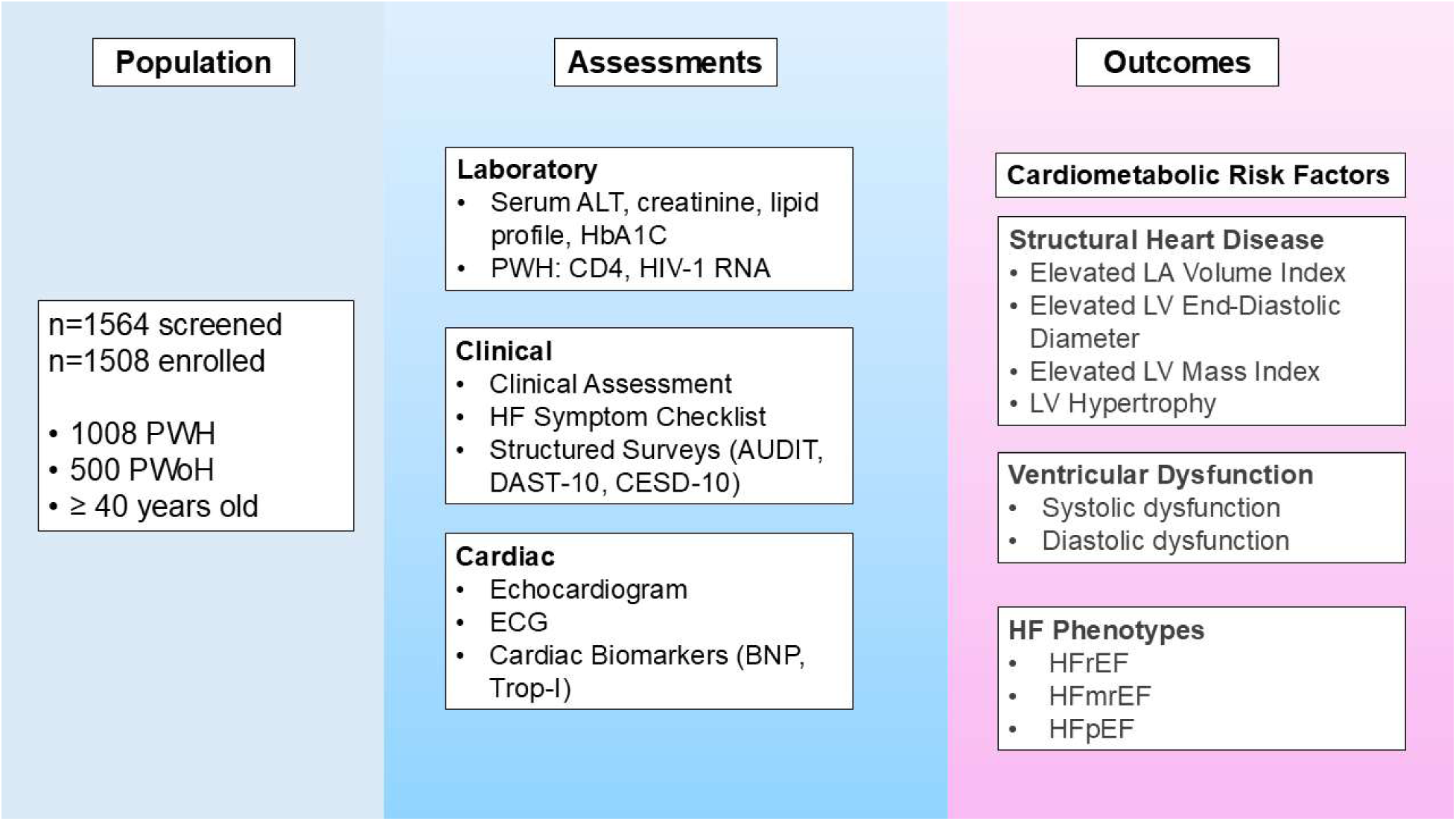
Study design schematic. ALT=alanine aminotransferase; AUDIT=Alcohol Use Disorders Identification Test; BNP=b-type natriuretic peptide; CESD-10=Center for Epidemiologic Studies Depression Scale-10; DAST-10=Drug Abuse Screen Test-10; ECG=electrocardiogram; HbA1C=hemoglobin A1C; HFmrEF=heart failure with mildly reduced ejection fraction; HFpEF=heart failure with preserved ejection fraction; HFrEF=heart failure with reduced ejection fraction; LA=left atrium; LV=left ventricle; PWH=people with HIV; PWoH=people without HIV; Trop-T=Troponin-I

Structured behavioral surveys were conducted including the Alcohol Use Disorders Identification Test (AUDIT), Drug Abuse Screening Test-10 (DAST-10), and Center for Epidemiological Studies Depression Scale-10 (CESD-10).

### Outcomes

Cardiometabolic risk factors of interest included hypertension (known diagnosis on medical chart review, use of antihypertensive therapy, or elevated blood pressure at recruitment [systolic BP>160 or diastolic BP > 100]), obesity (BMI ≥30 kg/m^2^), diabetes mellitus (known diagnosis, use of antidiabetic therapy, or serum hemoglobin A1C level ≥6.5% at recruitment), increased low density lipoprotein cholesterol (LDL-c) (LDL-c level ≥130 mg/dL [3.36 mmol/L] or use of lipid lowering therapy), chronic kidney disease (CKD stage IIIa or higher based on estimated glomerular filtration rate), and coronary artery disease (known diagnosis). Depressive symptoms were defined by a score of ≥10 on the CESD-10 scale, hazardous or harmful alcohol use defined by a score of ≥8 on the AUDIT scale, and moderate or greater illicit substance abuse by a score of ≥3 on the DAST-10 scale.

The American Heart Association (AHA) progressive HF stages were characterized, consisting of: Stage A, defined by the presence of cardiometabolic risk factors for HF without evidence of structural or functional cardiac abnormalities; Stage B, defined by ECHO evidence of either LV systolic or diastolic dysfunction; and Stage C (clinical HF), which included people with HF symptoms (from 5-question checklist) or BNP level of ≥35 pg/mL, and echo abnormalities from stage B. No participants in our sample were classified as advanced HF (Stage D) - these patients are immediately referred to a specialty cardiology unit and would not typically receive care at the recruitment site.

The heart disease outcomes of interest include structural cardiac abnormalities, ventricular dysfunction, and clinical HF definitions (detailed criteria in **Table 1**). Standardized published ECHO criteria were used to define the cardiac structural and functional abnormalities, and increased LV filling pressures.^14^

**Table 1:**
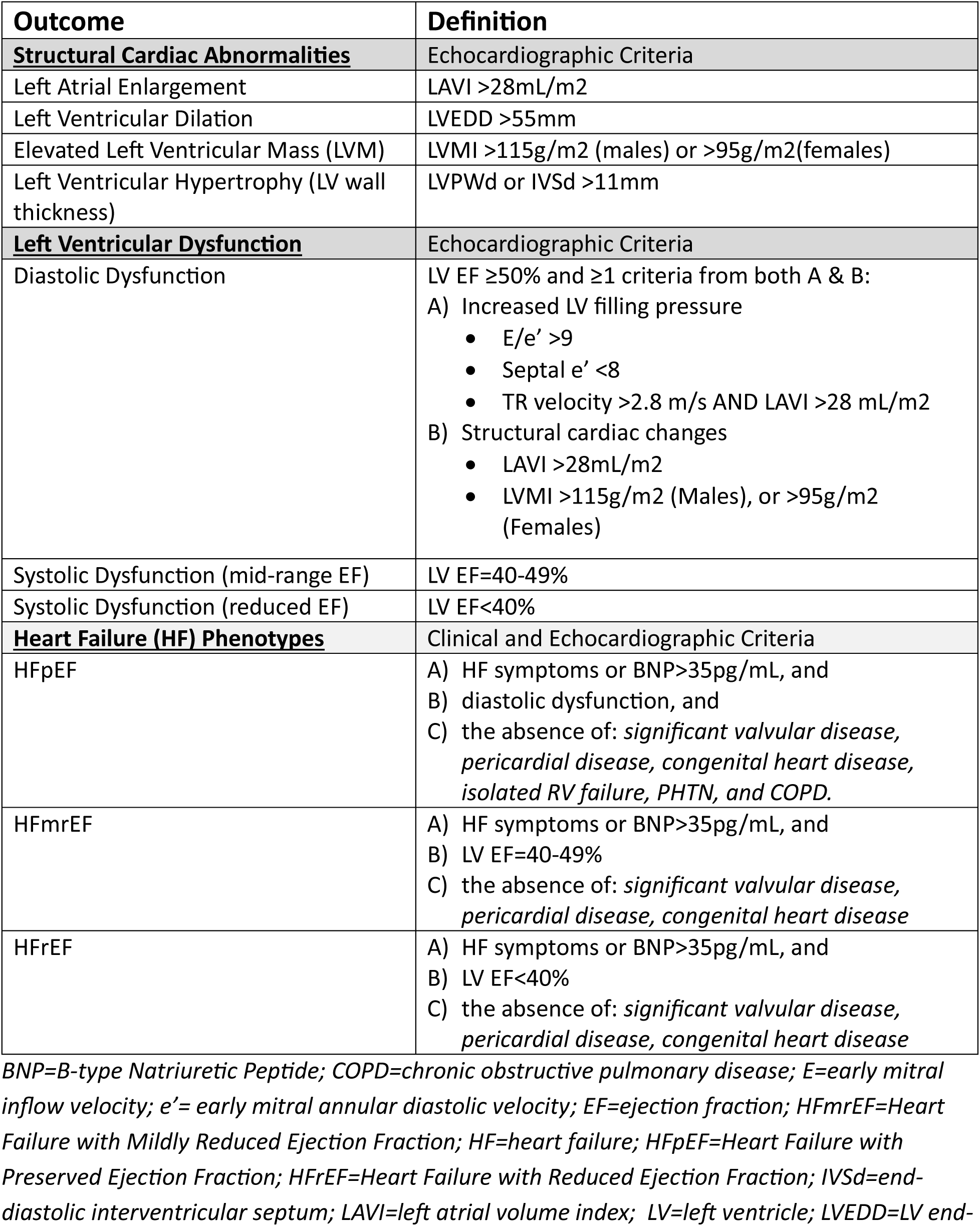
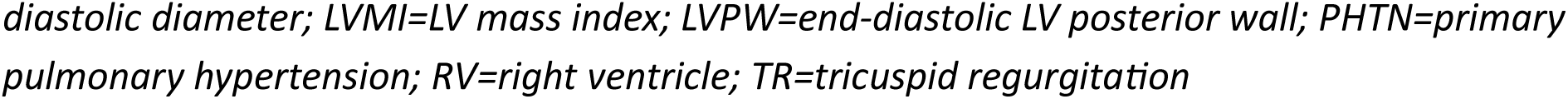
Outcome Definitions for Structural Cardiac Abnormalities, Ventricular Dysfunction, and Heart Failure Phenotypes.

| Outcome | Definition |
| --- | --- |
| **<u>Structural Cardiac Abnormalities</u>** | Echocardiographic Criteria |
| Left Atrial Enlargement | LAVI >28mL/m<sup>2</sup> |
| Left Ventricular Dilation | LVEDD >55mm |
| Elevated Left Ventricular Mass (LVM) | LVMI >115g/m<sup>2</sup> (males) or >95g/m<sup>2</sup>(females) |
| Left Ventricular Hypertrophy (LV wall thickness) | LVPWd or IVSd >11mm |
| **<u>Left Ventricular Dysfunction</u>** | Echocardiographic Criteria |
| Diastolic Dysfunction | LV EF ≥50% and ≥1 criteria from both A & B:<br>A) Increased LV filling pressure<br>E/e' >9<br>Septal e' <8<br>TR velocity >2.8 m/s AND LAVI >28 mL/m<sup>2</sup><br>B) Structural cardiac changes<br>LAVI >28mL/m<sup>2</sup><br>LVMI >115g/m<sup>2</sup> (Males), or >95g/m<sup>2</sup> (Females) |
| Systolic Dysfunction (mid-range EF) | LV EF=40-49% |
| Systolic Dysfunction (reduced EF) | LV EF<40% |
| **<u>Heart Failure (HF) Phenotypes</u>** | Clinical and Echocardiographic Criteria |
| HFpEF | A) HF symptoms or BNP>35pg/mL, and<br>B) diastolic dysfunction, and<br>C) the absence of: *significant valvular disease, pericardial disease, congenital heart disease, isolated RV failure, PHTN, and COPD.* |
| HFmrEF | A) HF symptoms or BNP>35pg/mL, and<br>B) LV EF=40-49%<br>C) the absence of: *significant valvular disease, pericardial disease, congenital heart disease* |
| HFrEF | A) HF symptoms or BNP>35pg/mL, and<br>B) LV EF<40%<br>C) the absence of: *significant valvular disease, pericardial disease, congenital heart disease* |

BNP=B-type Natriuretic Peptide; COPD=chronic obstructive pulmonary disease; E=early mitral inflow velocity; e' = early mitral annular diastolic velocity; EF=ejection fraction; HFmrEF=Heart Failure with Mildly Reduced Ejection Fraction; HF=heart failure; HFpEF=Heart Failure with Preserved Ejection Fraction; HFrEF=Heart Failure with Reduced Ejection Fraction; IVSd=end-diastolic interventricular septum; LAVI=left atrial volume index; LV=left ventricle; LVEDD=LV end-
*diastolic diameter; LVMI=LV mass index; LVPW=end-diastolic LV posterior wall; PHTN=primary* *pulmonary hypertension; RV=right ventricle; TR=tricuspid regurgitation*

### Statistical Analysis

At an estimated prevalence of HF and ventricular dysfunction in the range of 5-30%, our sample size was calculated at 1000 PWH and 500 PWoH to estimate prevalence with a margin of error of < 4 percentage points and yield 80% power to detect a difference of 7 percentage points by HIV status.

Participant characteristics, ECHO parameters and additional outcomes were summarized by median (Q1-Q3) for continuous variables and proportion (%) for categorical variables. Outcomes were compared between groups using univariable and multivariable logistic regression models including the following hypothesized confounding factors: age, sex, hypertension, smoking, obesity, diabetes, elevated LDL-c, and harmful/hazardous alcohol use. Comparisons by HIV status were then evaluated separately for men and women. Models also considered multiplicative interaction terms for HIV-by-sex and HIV-by-age.

## Results

From September 2022 to July 2025, 1508 participants were enrolled (1008 PWH and 500 PWoH). Only participants with complete echo data were included for analysis. Participant characteristics are presented in **Table 2** and HIV-specific factors in **Table 3**. The median age of all participants was 48 years, 77.3% were female, and the frequencies of hypertension (37.7%) and obesity (41.5%) were high, while frequencies of previously diagnosed coronary artery disease (0.4%) and chronic kidney disease (0.3%) were low. Among PWH, the majority (73.7%) had been living with HIV for over 10 years, 96.4% had a suppressed (<200 copies/mL) HIV RNA level at enrollment, and 37.9% had a nadir CD4 count <200 cells/μL. The median current CD4 count was 588 cells/μL, and the frequency of previous TB disease was 3-fold higher among PWH vs. PWoH (48.9% vs 16.0%, respectively).

**Table 2:**
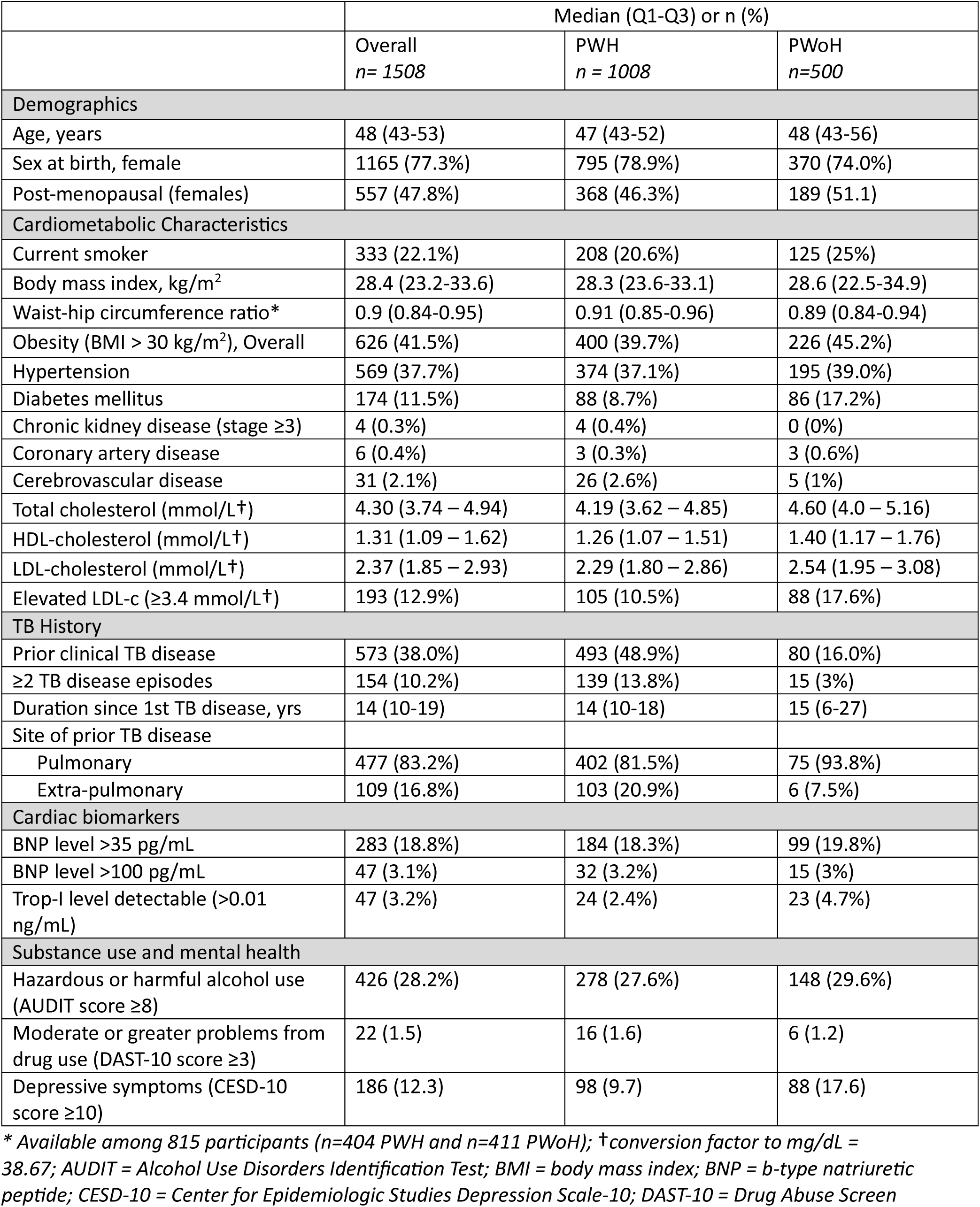
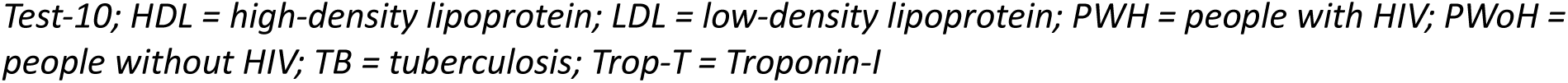
Participant characteristics.

|  | Median (Q1-Q3) or n (%) |  |  |
| --- | --- | --- | --- |
|  | Overall<br>*n* = 1508 | PWH<br>*n* = 1008 | PWoH<br>*n* = 500 |
| **Demographics** |  |  |  |
| Age, years | 48 (43-53) | 47 (43-52) | 48 (43-56) |
| Sex at birth, female | 1165 (77.3%) | 795 (78.9%) | 370 (74.0%) |
| Post-menopausal (females) | 557 (47.8%) | 368 (46.3%) | 189 (51.1) |
| **Cardiometabolic Characteristics** |  |  |  |
| Current smoker | 333 (22.1%) | 208 (20.6%) | 125 (25%) |
| Body mass index, kg/m<sup>2</sup> | 28.4 (23.2-33.6) | 28.3 (23.6-33.1) | 28.6 (22.5-34.9) |
| Waist-hip circumference ratio* | 0.9 (0.84-0.95) | 0.91 (0.85-0.96) | 0.89 (0.84-0.94) |
| Obesity (BMI > 30 kg/m<sup>2</sup>), Overall | 626 (41.5%) | 400 (39.7%) | 226 (45.2%) |
| Hypertension | 569 (37.7%) | 374 (37.1%) | 195 (39.0%) |
| Diabetes mellitus | 174 (11.5%) | 88 (8.7%) | 86 (17.2%) |
| Chronic kidney disease (stage ≥3) | 4 (0.3%) | 4 (0.4%) | 0 (0%) |
| Coronary artery disease | 6 (0.4%) | 3 (0.3%) | 3 (0.6%) |
| Cerebrovascular disease | 31 (2.1%) | 26 (2.6%) | 5 (1%) |
| Total cholesterol (mmol/L†) | 4.30 (3.74 – 4.94) | 4.19 (3.62 – 4.85) | 4.60 (4.0 – 5.16) |
| HDL-cholesterol (mmol/L†) | 1.31 (1.09 – 1.62) | 1.26 (1.07 – 1.51) | 1.40 (1.17 – 1.76) |
| LDL-cholesterol (mmol/L†) | 2.37 (1.85 – 2.93) | 2.29 (1.80 – 2.86) | 2.54 (1.95 – 3.08) |
| Elevated LDL-c (≥3.4 mmol/L†) | 193 (12.9%) | 105 (10.5%) | 88 (17.6%) |
| **TB History** |  |  |  |
| Prior clinical TB disease | 573 (38.0%) | 493 (48.9%) | 80 (16.0%) |
| ≥2 TB disease episodes | 154 (10.2%) | 139 (13.8%) | 15 (3%) |
| Duration since 1st TB disease, yrs | 14 (10-19) | 14 (10-18) | 15 (6-27) |
| Site of prior TB disease |  |  |  |
| Pulmonary | 477 (83.2%) | 402 (81.5%) | 75 (93.8%) |
| Extra-pulmonary | 109 (16.8%) | 103 (20.9%) | 6 (7.5%) |
| **Cardiac biomarkers** |  |  |  |
| BNP level >35 pg/mL | 283 (18.8%) | 184 (18.3%) | 99 (19.8%) |
| BNP level >100 pg/mL | 47 (3.1%) | 32 (3.2%) | 15 (3%) |
| Trop-I level detectable (>0.01 ng/mL) | 47 (3.2%) | 24 (2.4%) | 23 (4.7%) |
| **Substance use and mental health** |  |  |  |
| Hazardous or harmful alcohol use (AUDIT score ≥8) | 426 (28.2%) | 278 (27.6%) | 148 (29.6%) |
| Moderate or greater problems from drug use (DAST-10 score ≥3) | 22 (1.5) | 16 (1.6) | 6 (1.2) |
| Depressive symptoms (CESD-10 score ≥10) | 186 (12.3) | 98 (9.7) | 88 (17.6) |

\* Available among 815 participants (*n*=404 PWH and *n*=411 PWoH); †conversion factor to mg/dL = 38.67; AUDIT = Alcohol Use Disorders Identification Test; BMI = body mass index; BNP = b-type natriuretic peptide; CESD-10 = Center for Epidemiologic Studies Depression Scale-10; DAST-10 = Drug Abuse Screen
*Test-10; HDL = high-density lipoprotein; LDL = low-density lipoprotein; PWH = people with HIV; PWoH =* *people without HIV; TB = tuberculosis; Trop-T = Troponin-I*

**Table 3:** HIV-specific participant characteristics.

|  | Median (Q1-Q3) or n (%) |
| --- | --- |
|  | PWH<br>*n=1008* |
| HIV diagnosis duration, years |  |
| <5 years | 74 (7.3%) |
| 5-10 years | 191 (19.0%) |
| >10 years | 742 (73.7%) |
| HIV diagnosis duration, years | 14.3 (10-18) |
| Duration since ART initiation, years | 12.0 (8-15) |
| HIV-1 RNA level <200 copies/mL | 972 (96.4%) |
| HIV-1 RNA level <200 copies/mL for ≥3 years | 680 (67.5%) |
| CD4 count | 587.5 (425-746) |
| Nadir CD4 count <200 cells/μL | 382 (37.9%) |
| Current ART use |  |
| dolutegravir | 959 (95.1%) |
| tenofovir disoproxil fumarate | 939 (93.2%) |
| lamivudine | 937 (96.5%) |
| abacavir | 41 (4.1%) |
| Prior ART exposure |  |
| efavirenz | 886 (89.9%) |
| emtricitabine | 865 (85.8%) |
| stavudine | 205 (20.3%) |
| zidovudine | 168 (16.7%) |
| ritonavir-boosted lopinavir | 81 (8%) |
| abacavir | 13 (1.3%) |

*ART = antiretroviral therapy; PWH = people with HIV; RNA=ribonucleic acid*

Overall, 65% of participants had at least one cardiometabolic risk factor. The frequency of hypertension was similar (37.2% in PWH vs 39% in PWoH) in both groups by intention, but PWoH more commonly had obesity (39.7% vs 45.2%), diabetes (8.7% vs 17.2%) or elevated LDL-c levels (10.4% vs 17.6%).

Classification of progressive AHA HF stages is shown in **Table 4**. Both groups had similar frequencies of participants without risk factors or evidence of structural heart disease (no AHA stage), though AHA Stage A was more common among PWoH (27.1% vs 36.8%; p<0.001). However, compared to PWoH, the proportion of AHA Stage B (structural or functional heart disease without symptoms) was significantly higher among PWH (38.9% vs. 32.0%; p=0.011), while AHA Stage C (clinical HF) (10.5% vs 9.0%; p=0.405) was not significantly different.

**Table 4:** Classification across Mutually Exclusive AHA Heart Failure Stages.

| AHA Heart Failure Stage | *n* per stage | PWH<br>*n*=1008 | PWoH<br>*n*=500 | *p*-value |
| --- | --- | --- | --- | --- |
| No stage | 348 | 237 (23.5%) | 111 (22.2%) | 0.614 |
| AHA Stage A (Risk factors* for heart failure with no evidence of disease) | 457 | 273 (27.1%) | 184 (36.8%) | <0.001 |
| AHA Stage B (Echo abnormalities† but asymptomatic and normal BNP) | 552 | 392 (38.9%) | 160 (32%) | 0.011 |
| AHA Stage C (LV dysfunction with HF symptoms or BNP≥35pg/mL) | 151 | 106 (10.5%) | 45 (9%) | 0.405 |

*Stages are mutually exclusive. \*Risk factors included diabetes, hypertension, elevated LDL-* *cholesterol (LDL-c≥ 3.362 mmol/L), obesity, chronic kidney disease stages 3-5, and coronary* *artery disease. †Echo abnormalities assessed include LA volume index, LV mass index, LV* *hypertrophy, LV end-diastolic diameter, systolic and diastolic dysfunction, and significant valvular* *disease. Stages are mutually exclusive. No participant was classified into Stage D. PWH=people* *with HIV; PWoH=people without HIV; AHA=American heart Association; BNP=B-type natriuretic* *peptide; LV=left ventricle*

Frequencies of cardiac structural abnormalities, LV dysfunction, and HF syndromes are reported in **Supplemental Table 1,** and the risk of these outcomes by HIV status is presented in **Figure 2**. When compared to PWoH, PWH were significantly less likely to have elevated LVEDD (5.1% in PWH vs 10.0% in PWoH; OR 0.50; 95% CI 0.33-0.76; p<0.01), and more likely to have increased LVMI (19.9% in PWH vs 11.6%; OR 2.14; 95% CI 1.54-3.02 p<0.001) and LV wall thickness (20.6% vs 7.6%; OR 3.76; 95% CI 2.60-5.58; p<0.001). Systolic dysfunction did not differ, but diastolic dysfunction was more frequent among PWH compared to PWoH (15.8% vs. 12.8%; OR 1.43; 95% CI 1.03-2.01; p=0.033) (**Supplemental Table 1**).

**Figure 2:**
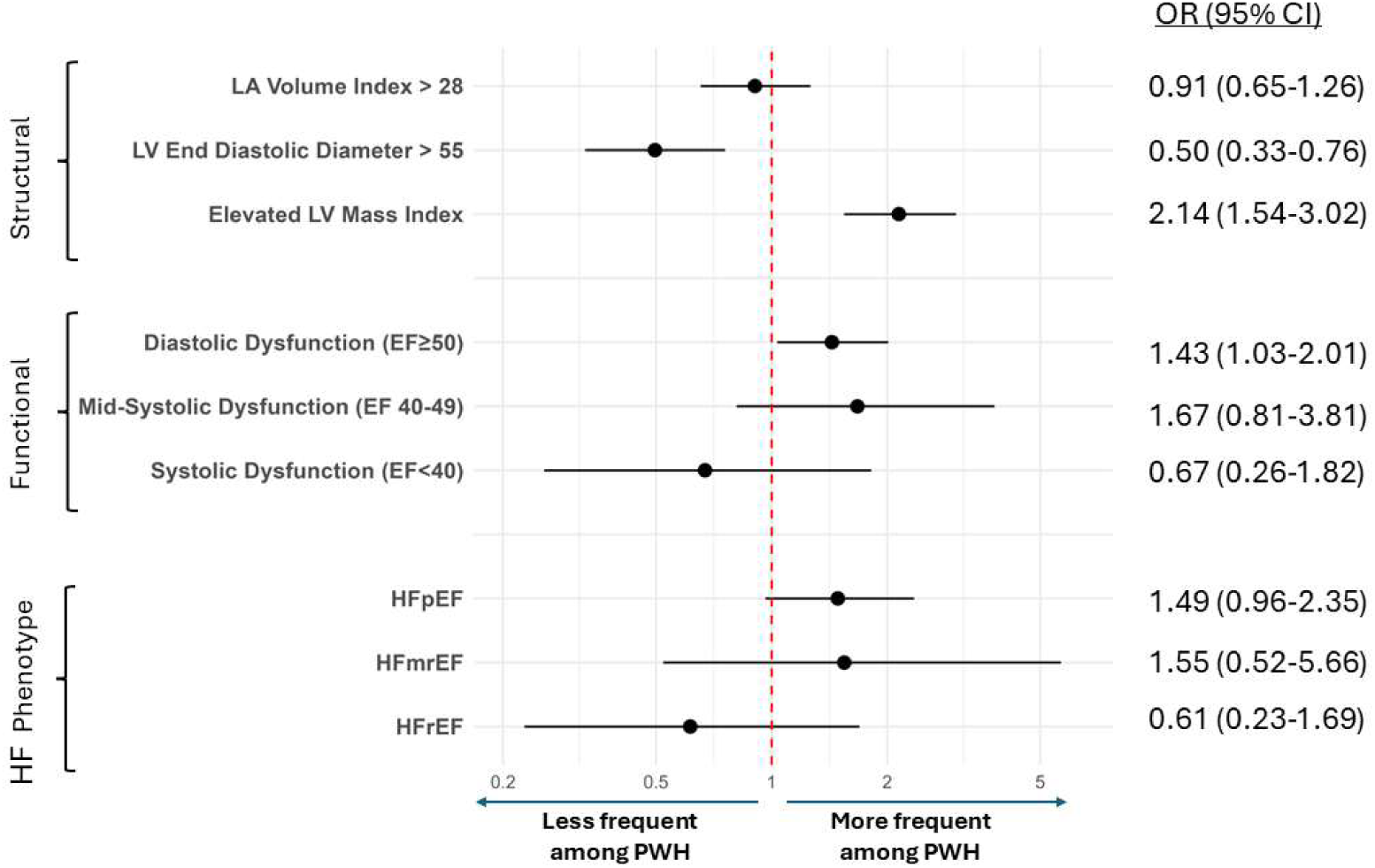
OR (95% CI) for myocardial abnormalities among people with vs without HIV. OR (95% CI) plotted for each cardiac variable comparing people with vs. without HIV (PWH, PWoH). OR adjusted for age, sex, diabetes, hypertension, elevated LDL-cholesterol (LDL-c ≥ 3.362 mmol/L), smoking and hazardous alcohol use. HF=heart failure; HFmrEF=Heart Failure with Mildly Reduced Ejection Fraction; HFpEF=Heart Failure with Preserved Ejection Fraction; HFrEF=Heart Failure with Reduced Ejection Fraction; LA=left atrium; LV=left ventricle.

Overall, HFpEF was the predominant HF phenotype (7.8%) while HFrEF was rare (1.1%). Compared to PWoH, HFpEF was more frequent among PWH (8.4% vs 6.6%) though this difference did not reach statistical significance in unadjusted (OR 1.30; 95% CI 0.87-2.0; P=0.21) or adjusted (OR 1.49; 95% CI 0.96-2.35; p=0.08) models. Other factors associated with HFpEF included older age, female sex, hypertension, and current or past smoking (**Table 5**). When considering HIV-specific factors among PWH, risk for HFpEF was associated with having a current CD4 count <200 cells/μL (OR 4.49; 95% CI 1.68-10.8; p<0.01; **Supplemental Table 2**). Exposure to protease inhibitor-containing ART regimens also had a large but non-significant point estimate for an association with HFpEF (OR 1.86; 95% CI 0.88-3.62; p=0.082). There was no association with duration of HIV infection, duration of ART use, or exposure to integrase strand transfer inhibitor-containing ART regimens. No TB-specific factors were significantly associated with HFpEF (**Supplemental Table 3**).

**Table 5:** Odds Ratios for HFpEF Risk in Multivariable Adjusted Model.

|  | Odds Ratio (95% CI) | p value |
| --- | --- | --- |
| HIV | 1.49 (0.96-2.35) | 0.080 |
| Age per 10-year increase | 1.82 (1.39-2.39) | <0.001 |
| Female sex | 2.34 (1.22-4.64) | 0.012 |
| Obesity | 1.53 (0.99-2.36) | 0.054 |
| Hypertension | 1.71 (1.12-2.61) | 0.012 |
| LDL-c ≥ 3.362 mmol/L | 0.87 (0.46-1.53) | 0.651 |
| Diabetes | 0.73 (0.39-1.31) | 0.319 |
| Smoking | 1.75 (1.0-2.99) | 0.043 |
| Hazardous alcohol use | 1.12 (0.66-1.86) | 0.667 |

*HFpEF risk adjusted for age, sex, obesity, diabetes, hypertension, elevated LDL-cholesterol (LDL-* *c ≥ 3.362 mmol/L), smoking, and hazardous or harmful alcohol use*

The potential influence of sex on HFpEF associations with HIV status was further examined, given substantial differences in frequencies of hypertension (27.1% vs 40.9%), obesity (6.7% vs 51.8%) and diabetes (5.0% vs 13.5%) between males and females, respectively (**Supplemental Tables 4 and 5**). When comparisons were restricted to females, the magnitude of increased risk among PWH was greater for LVMI (OR 2.39; 95% CI 1.65-3.51; p<0.001), LV wall thickness (OR 4.47; 95% CI 2.84-7.33; p<0.001), and diastolic dysfunction (OR 1.64; 95% CI 1.13-2.42; p=0.011), when compared to estimates among men or among all participants (**Figure 3** and **Supplemental Tables 6 and 7**). Risk for HFpEF was now significantly higher for women with HIV compared to women without HIV (OR 1.88; 95% CI 1.14-3.20; p=0.016). Further, there was suggestion of heterogeneity in the effect of HIV on risk for HFpEF between females and males (p-value for the HIV-by-sex interaction = 0.051). HIV-by-age (p=0.953) and HIV-by-menopausal status (p=0.973) interaction terms were not significant.

**Figure 3:**
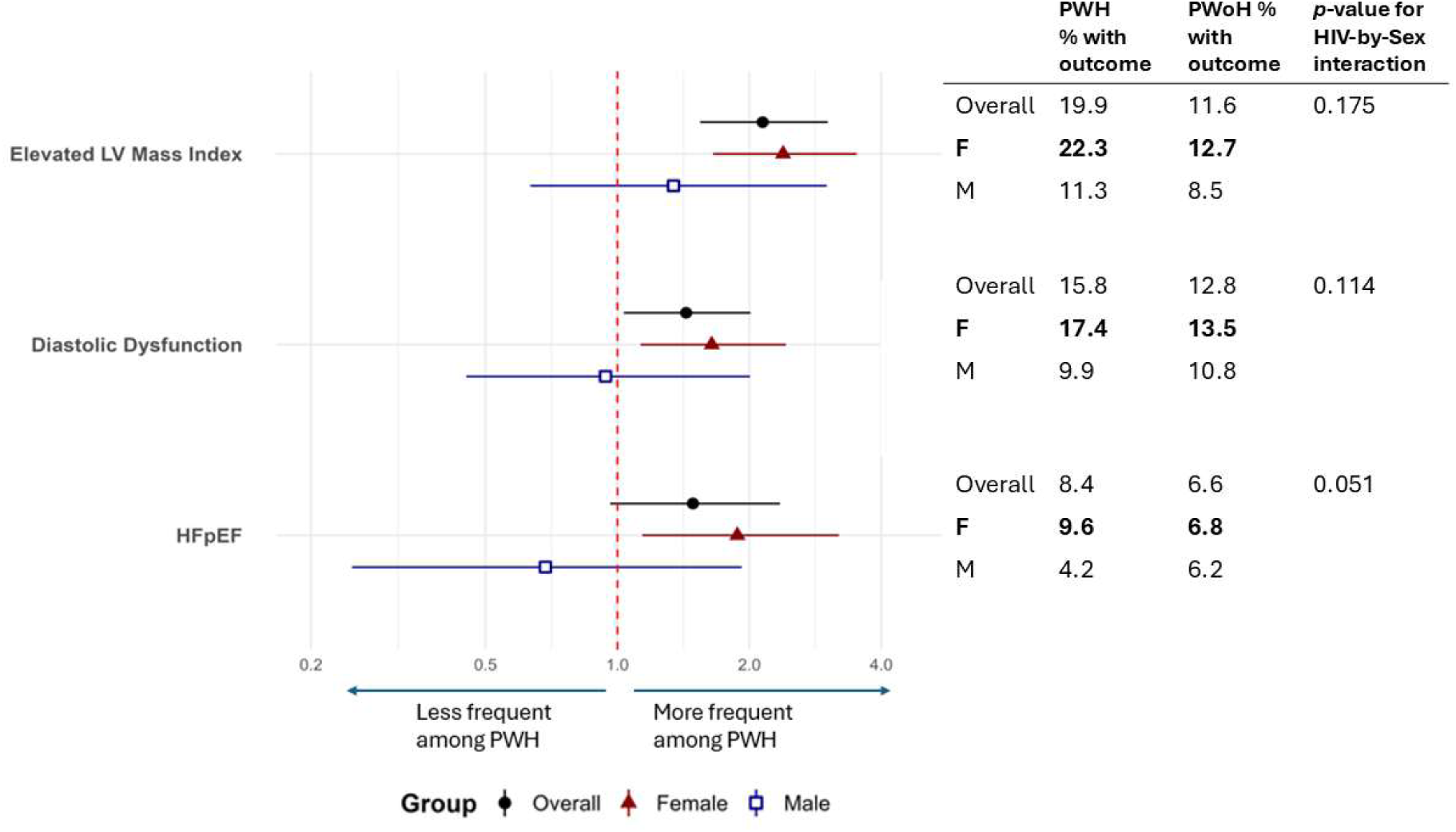
Risk for myocardial abnormalities by HIV status and sex. OR (95% CI) plotted for each cardiac variable comparing people with vs. without HIV (PWH, PWoH), separated by sex. OR adjusted for age, diabetes, hypertension, elevated LDL-cholesterol (LDL-c ≥3.362 mmol/L), smoking and hazardous alcohol use.

## Discussion

We described the frequency of structural and functional cardiac abnormalities, and undiagnosed HF phenotypes, among a population of ART-treated PWH and PWoH in a front-line clinical setting within Khayelitsha, South Africa (SA). There are four primary findings: (1) Traditional cardiometabolic risk factors (e.g., hypertension and obesity) and subclinical cardiac abnormalities (e.g., LV wall thickness) characterized by ECHO were very common among both persons with and without HIV; (2) The overall frequency of undiagnosed AHA HF stages B and C was unexpectedly high, and amongst those with AHA HF stage C, HFpEF was the predominant HF phenotype while HFrEF was rare; (3) Compared to PWoH, PWH had a higher frequency of ECHO-determined LV wall thickness and increased LV mass, as well as diastolic dysfunction, both of which often precede and contribute to clinical HFpEF risk; and (4) in this community, the overall frequencies of structural and functional cardiac abnormalities and risk for undiagnosed HFpEF were significantly higher among women with HIV versus women without HIV, independent of cardiometabolic risk factors.

While there is wide acceptance that CVD risk is increased among PWH, most of the evidence to date focuses on coronary atherosclerosis and its associated ischemia-induced clinical manifestations.^15^ Accelerated atherogenesis among PWH is multifactorial, but is due, in part, to high prevalence of cardiometabolic risk factors, adverse effects and toxicities of antiretroviral drugs, and persistent systemic inflammation and immune activation despite plasma viral suppression with ART.^16–19^ However, the degree to which these global North derived data are generalizable to regions such as SSA, where epidemiologic data for CVD in the general population suggest a predominance of non-ischemic heart disease that is less influenced by coronary atherogenesis per se, is unclear,.^6–8,20^

To our knowledge, this is the first SA based study to describe the prevalence of HF and HF phenotypes among an otherwise healthy community population including PWH receiving effective ART treatment. The high prevalence of traditional cardiometabolic risk factors among PWH in this study was consistent with data from other SSA cohorts (comparisons presented in **Supplemental Table 8)**. However, compared to studies conducted in the global North (**Supplemental Table 9**), PWH in PHIZA had less frequent elevations in non-HDL cholesterol and more frequent obesity and hypertension. Both obesity and hypertension are not only risk factors for heart failure but may be causally related, ^21–23^ potentially contributing to the high prevalence of undiagnosed HFpEF overall in PHIZA.

Data from the global North have also shown that HIV is associated with increased risk for both HFrEF and HFpEF phenotypes. ^24^ HIV-associated HF in the pre-ART era, with advanced untreated HIV/AIDS, was typically characterized by myocarditis and myonecrosis leading to dilated cardiomyopathy with a reduced LV systolic function.^25^ With the widespread use of ART and associated viral suppression and immune recovery, HIV-associated dilated cardiomyopathy is less common in contemporary settings. Recent ECHO data among ART-treated PWH with high CD4 counts suggest a higher burden of diastolic dysfunction.^26,27^ In PHIZA, HFpEF was the predominant HF phenotype regardless of HIV status. Further, when compared to controls, PWH had a 171% higher risk of having increased LV wall thickness, 71% higher risk of having increased LV mass, and 23% higher risk for diastolic dysfunction, all precursors to HFpEF.

Comparisons of HF phenotypes with other data from SSA are difficult due to the lack of population-based studies with detailed echocardiographic assessments. Prior epidemiologic data from SSA support a similar predominance of non-ischemic CVD manifestations including HF phenotypes, but these data are either outdated (e.g., from the pre-ART era among those with advanced AIDS) or have design limitations (such as small sample sizes, mixed reporting of ART use, or reporting surrogate CVD markers without clinical CVD phenotypes).^10,12,28^ For example, the Heart of Soweto^20^ (which assessed 4810 patients of which 9.7% were PWH) was designed to determine the etiology of CVD in a peri-urban Johannesburg (SA) community with a similar demographic and socioeconomic background to Khayelitsha and similar burden of HIV.^29^ Non-ischemic HF was the most common CVD manifestation (44%), while ASCVD complications, such as ischemic heart disease (2.7%) and cerebrovascular disease (3.5%) were infrequent. Further analyses from the Heart of Soweto study, limited to only participants with HIV, found ASCVD-related admissions in only 2.4%.^28^ Consistent with this, emerging CVD epidemiology from the general population in SSA report the most common causes of HF as hypertensive heart disease, cardiomyopathy, and rheumatic heart disease^6^, with limited contribution from coronary artery disease.^5,6,8^ These findings highlight the importance of contemporary studies from the global South like PHIZA, and the potential hazard of relying on information derived from populations defined by a different spectrum of risk factors.

Possible mechanisms contributing to risk for HFpEF among PWH receiving effective ART include HIV-related systemic inflammation and structural changes characterized by excess myocardial fibrosis and steatosis.^15^ Evidence of HIV-associated myocardial fibrosis and steatosis has now been reported in numerous studies utilizing cardiac magnetic resonance imaging, as well as histopathology from a sentinel autopsy study.^15,30–33^ Among PWH, myocardial fibrosis has also been associated with circulating proinflammatory monocytes and a plasma proteomic signature enriched for T cell activation and TNF signaling, suggesting a possible immune mechanism for cardiac inflammation.^32,34,35^ Finally, multiple studies from the general population have reported associations for diastolic dysfunction and risk for HFpEF with systemic inflammation, endothelial dysfunction, and myocardial steatosis, all of which are accentuated among PWH.^17,21,36–38^

Our findings build on existing literature by identifying that women with HIV may have a higher frequency of undiagnosed HFpEF compared to women without HIV, whereas this effect may not be present among men (**Figure 3; Supplemental Tables 6 and 7**). In addition to having higher frequencies of hypertension and obesity, both key risk factors for HFpEF, the association between HIV status and increased LV mass, LV hypertrophy, and diastolic dysfunction was more prominent among women. (**Figure 3; Supplemental Tables 4 and 5**). This is consistent with a recent analysis of the global REPRIEVE trial which reported that incident HF was higher in women and those with hypertension and obesity.^39^ Furthermore, outside of HICs, incident HF was reported more frequently in SSA than other regions in REPRIEVE.^39^ Among a study of out of hospital cardiac arrest in the U.S., PWH were at increased risk for sudden cardiac death when compared to persons without HIV in the same community, and this effect was greater among women than men.^33^ Finally, the observation that the relative increase in CVD risk from HIV is greater among women, compared to men, has also been repeatedly shown for ASCVD events.^40,41^ Some have posited that sex differences in HIV-related CVD may be due to underlying differences in immune activation, which could have implications for both ischemic and non-ischemic CVD manifestations.^42^ Future research is needed to better characterize the potential interaction between HIV disease, obesity, and female sex, as well as the associated mechanisms contributing to LV wall thickness, diastolic dysfunction and clinical risk from HFpEF.

The clinical syndrome of HFpEF has been characterized as the end result of cardiac insults and stress from numerous heterogeneous comorbidities.^21^ Recently, a novel unifying biologic framework has been proposed that describes the pathogenesis of HFpEF arising from dysfunctional adipose tissue rather than as a primary disorder of cardiomyocytes.^22^ This framework has been referred to as the “adipokine hypothesis”, which states that an expansion of altered visceral adipose tissue leads to an increased and altered profile of adipokine release into the circulating blood volume, which over time contributes to systemic inflammation, plasma volume expansion and cardiac hypertrophy and fibrosis.^22^ In this context, women have increased adipokines and related inflammatory markers when compared to men.^43^ Further, women are at a two-fold increased risk for HFpEF when compared to men, and obesity poses greater risk for HFpEF among women when compared to men.^44^ Applying the ’adipokine hypothesis’ to PWH in the current era, it is notable that chronic inflammation and adiposity may both be more prominent among women when compared to men with HIV.^42,45^ Specific to ART treatment, weight gain has been associated with integrase strand transfer inhibitor-based regimens, with the greatest weight gain reported among black women.^45^ In summary, our findings suggest that ART-treated HIV disease may amplify key components of the ‘adipokine hypothesis’, namely inflammation and visceral adiposity, thereby accentuating the pathogenesis of HFpEF particularly among women with HIV.

This study had several important limitations. The cross-sectional design inherently limits causal inference and may also include unmeasured confounding. The clinical and laboratory assessments were limited within the frontline clinic research setting, which prevented exploration of immunologic and inflammatory pathways that may be contributing to the cardiac outcomes described. Finally, the study population age was primarily between 40 to 60 years old, but this reflects an age range at higher risk for CVD where prevention strategies are also still useful. The study population was predominantly women, reflective of the epidemiology within SA, with approximately two thirds of adult PWH in SA being women^29^; nonetheless, this limited our ability to make comparisons restricted to men only. Additionally, comparisons between men and women were potentially confounded by meaningful differences in the profile of cardiometabolic risk factors.

## Conclusion

In a peri-urban South African (SA) community with low socioeconomic status and a high prevalence of HIV, TB and cardiometabolic risk factors, undiagnosed HFpEF was common and was the predominant HF phenotype. HIV was associated with increased risk for subclinical structural and functional cardiac abnormalities including greater LV mass, LV wall thickness, and diastolic dysfunction, which all contribute to HFpEF risk. These findings were more prominent among women with HIV when compared to women without HIV. In this context, PWH in SA, particularly women, have an increased risk for HF that is characterized by thick, stiff myocardium, independent of cardiometabolic risk factors. This large community-based study adds to increasing evidence suggesting that in SSA, where the prevalence of HIV is high and the burden of ASCVD is relatively low,^5^ the absolute excess CVD risk due to HIV manifests most frequently as non-ischemic HF and HFpEF. Further research should focus on the potential interaction and interconnectedness of HIV disease, female sex and obesity along with their influence on HFpEF pathogenesis.

## Data Availability

A dataset will be made available upon request.

## Non-standard Abbreviations and Acronyms

AHA: American heart Association
ART: Antiretroviral therapy
ASCVD: Atherosclerotic cardiovascular disease
AUDIT: Alcohol Use Disorders Identification Test
BNP: B-type natriuretic peptide
CESD: Center for Epidemiological Studies Depression Scale-10
CVD: Cardiovascular disease
DAST-10: Drug Abuse Screening Test-10
DD: Diastolic dysfunction
ECHO: Echocardiogram
EF: Ejection fraction
HF: Heart failure
HFmrEF: Heart failure with mildly reduced ejection fraction
HFpEF: Heart failure with preserved ejection fraction
HFrEF: Heart failure with reduced ejection fraction
HIC: High income countries
LDL: Low density lipoprotein
LV: Left ventricle
LVMI: Left ventricle mass index
PWH: People with HIV
PWoH: People without HIV
SA: South Africa
SSA: sub-Saharan Africa

## Acknowledgements

We thank all our participants and the research staff at CIDRI-Africa and Hennepin Healthcare Research Institute for their contributions to the success of this study.

## Supplemental Material

- Supplemental Material including Supplemental Methods
- Tables S1–S11
- Figure S1
- References 46-50

## References

1. AIDS, crisis and the power to transform: UNAIDS Global AIDS Update 2025. Geneva: Joint United Nations Programme on HIV/AIDS; 2025. Licence: CC BY-NC-SA 3.0 IGO. https://www.unaids.org/en/resources/documents/2025/global-aids-update

2. Marcus JL, Leyden WA, Alexeeff SE, Anderson AN, Hechter RC, Hu H, Lam JO, Towner WJ, Yuan Q, Horberg MA, et al. Comparison of Overall and Comorbidity-Free Life Expectancy Between Insured Adults With and Without HIV Infection, 2000-2016. JAMA Netw. Open. 2020;3:e207954.

3. Shah ASV, Stelzle D, Lee KK, Beck EJ, Alam S, Clifford S, Longenecker CT, Strachan F, Bagchi S, Whiteley W, et al. Global Burden of Atherosclerotic Cardiovascular Disease in People Living With HIV. Circulation. 2018;138:1100–1112.

4. Ntsekhe M. Low Prevalence of Coronary Artery Disease Among Ugandans With and Without HIV: Local Anomaly or Regional Reality? Ann. Intern. Med. 2025;178:594–595.

5. Siedner MJ, Ghoshhajra B, Erem G, Nassanga R, Randhawa M, Ochieng A, Acan M, Lu MT, Thondapu V, Takigami A, et al. Epidemiology of Coronary Atherosclerosis Among People Living With HIV in Uganda. Ann. Intern. Med. 2025;178:468–478.

6. Yuyun MF, Sliwa K, Kengne AP, Mocumbi AO, Bukhman G. Cardiovascular diseases in sub-saharan Africa compared to high-income countries: An epidemiological perspective. Glob. Heart. 2020;15.

7. Keates AK, Mocumbi AO, Ntsekhe M, Sliwa K, Stewart S. Cardiovascular disease in Africa: Epidemiological profile and challenges. Nat. Rev. Cardiol. 2017;14:273–293.

8. Stark BA, DeCleene NK, Desai EC, Hsu JM, Johnson CO, Lara-Castor L, LeGrand KE, A PB, Aalipour MA, Aalruz H, et al. Global, Regional, and National Burden of Cardiovascular Diseases and Risk Factors in 204 Countries and Territories, 1990-2023. JACC. 2025;86:2167–2243.

9. Okello S, Amir A, Bloomfield GS, Kentoffio K, Lugobe HM, Reynolds Z, Magodoro IM, North CM, Okello E, Peck R, et al. Prevention of cardiovascular disease among people living with HIV in sub-Saharan Africa. Prog. Cardiovasc. Dis. 2020;63:149–159.

10. Hyle EP, Mayosi BM, Middelkoop K, Mosepele M, Martey EB, Walensky RP, Bekker LG, Triant VA. The association between HIV and atherosclerotic cardiovascular disease in sub-Saharan Africa: A systematic review. BMC Public Health. 2017;17.

11. Grinspoon SK, Fitch K V., Zanni M V., Fichtenbaum CJ, Umbleja T, Aberg JA, Overton ET, Malvestutto CD, Bloomfield GS, Currier JS, et al. Pitavastatin to Prevent Cardiovascular Disease in HIV Infection. New England Journal of Medicine. 2023;389:687–699.

12. Ntusi NAB, Ntsekhe M. Human immunodeficiency virus-associated heart failure in sub-Saharan Africa: evolution in the epidemiology, pathophysiology, and clinical manifestations in the antiretroviral era. ESC Heart Fail. 2016;3:158–167.

13. Critchley JA, Limb ES, Khakharia A, Carey IM, Auld SC, De Wilde S, Harris T, Phillips LS, Cook DG, Rhee MK, et al. Tuberculosis and Increased Incidence of Cardiovascular Disease: Cohort Study Using United States and United Kingdom Health Records. Clinical Infectious Diseases. 2025;80:271–279.

14. McDonagh TA, Metra M, Adamo M, Baumbach A, Böhm M, Burri H, Čelutkiene J, Chioncel O, Cleland JGF, Coats AJS, et al. 2021 ESC Guidelines for the diagnosis and treatment of acute and chronic heart failure. Eur. Heart J. 2021;42:3599–3726.

15. Ntsekhe M, Baker J V. Cardiovascular Disease among Persons Living with HIV: New Insights into Pathogenesis and Clinical Manifestations in a Global Context. Circulation. 2023;147:83–100.

16. Althoff KN, Gebo KA, Moore RD, Boyd CM, Justice AC, Wong C, Lucas GM, Klein MB, Kitahata MM, Crane H, et al. Contributions of traditional and HIV-related risk factors on non-AIDS-defining cancer, myocardial infarction, and end-stage liver and renal diseases in adults with HIV in the USA and Canada: a collaboration of cohort studies. Lancet HIV. 2019;6:e93–e104.

17. Grund B, Baker J V., Deeks SG, Wolfson J, Wentworth D, Cozzi-Lepri A, Cohen CJ, Phillips A, Lundgren JD, Neaton JD. Relevance of interleukin-6 and D-dimer for serious non-AIDS morbidity and death among HIV-positive adults on suppressive antiretroviral therapy. PLoS One. 2016;11.

18. Jaschinski N, Greenberg L, Neesgaard B, Miró JM, Grabmeier-Pfistershammer K, Wandeler G, Smith C, De Wit S, Wit F, Pelchen-Matthews A, et al. Recent abacavir use and incident cardiovascular disease in contemporary-treated people with HIV. AIDS. 2023;37:467–475.

19. Neesgaard B, Greenberg L, Miró JM, Grabmeier-Pfistershammer K, Wandeler G, Smith C, De Wit S, Wit F, Pelchen-Matthews A, Mussini C, et al. Associations between integrase strand-transfer inhibitors and cardiovascular disease in people living with HIV: a multicentre prospective study from the RESPOND cohort consortium. Lancet HIV. 2022;9:e474–e485.

20. Sliwa K, Wilkinson D, Hansen C, Ntyintyane L, Tibazarwa K, Becker A, Stewart S. Spectrum of heart disease and risk factors in a black urban population in South Africa (the Heart of Soweto Study): a cohort study. The Lancet. 2008;371:915–922.

21. Paulus WJ, Tschöpe C. A novel paradigm for heart failure with preserved ejection fraction: Comorbidities drive myocardial dysfunction and remodeling through coronary microvascular endothelial inflammation. J. Am. Coll. Cardiol. 2013;62:263–271.

22. Packer M. The Adipokine Hypothesis of Heart Failure With a Preserved Ejection Fraction: A Novel Framework to Explain Pathogenesis and Guide Treatment. J. Am. Coll. Cardiol. 2025;86:1269–1373.

23. Peikert A, Vaduganathan M, Claggett BL, Kulac IJ, Litwin S, Zile M, Desai AS, Jhund PS, Butt JH, Lam CSP, et al. Near-universal prevalence of central adiposity in heart failure with preserved ejection fraction: the PARAGON-HF trial. Eur. Heart J. 2025;46:2372–2390.

24. Freiberg MS, Chang CCH, Skanderson M, Patterson O V., DuVall SL, Brandt CA, So-Armah KA, Vasan RS, Oursler KA, Gottdiener J, et al. Association between HIV infection and the risk of heart failure with reduced ejection fraction and preserved ejection fraction in the antiretroviral therapy era: Results from the veterans aging cohort study. JAMA Cardiol. 2017;2:536–546.

25. Himelman RB, Chung WS, Chernoff DN, Schiller NB, Hollander H. Cardiac manifestations of human immunodeficiency virus infection: A two-dimensional echocardiographic study. J. Am. Coll. Cardiol. 1989;13:1030–1036.

26. Mondy KE, Gottdiener J, Overton ET, Henry K, Bush T, Conley L, Hammer J, Carpenter CC, Kojic E, Patel P, et al. High prevalence of echocardiographic abnormalities among HIV-infected persons in the era of highly active antiretroviral therapy. Clinical Infectious Diseases. 2011;52:378–386.

27. Cerrato E, D’Ascenzo F, Biondi-Zoccai G, Calcagno A, Frea S, Grosso Marra W, Castagno D, Omedè P, Quadri G, Sciuto F, et al. Cardiac dysfunction in pauci symptomatic human immunodeficiency virus patients: A meta-analysis in the highly active antiretroviral therapy era. Eur. Heart J. 2013;34:1432–1436.

28. Sliwa K, Carrington MJ, Becker A, Thienemann F, Ntsekhe M, Stewart S. Contribution of the human immunodeficiency virus/acquired immunodeficiency syndrome epidemic to de novo presentations of heart disease in the Heart of Soweto Study cohort. Eur. Heart J. 2012;33:866–874.

29. Kim H, Tanser F, Tomita A, Vandormael A, Cuadros DF. Beyond HIV prevalence: Identifying people living with HIV within underserved areas in South Africa. BMJ Glob. Health. 2021;6.

30. Ntusi N, O’Dwyer E, Dorrell L, Wainwright E, Piechnik S, Clutton G, Hancock G, Ferreira V, Cox P, Badri M, et al. HIV-1-Related Cardiovascular Disease Is Associated with Chronic Inflammation, Frequent Pericardial Effusions, and Probable Myocardial Edema. Circ. Cardiovasc. Imaging. 2016;9.

31. Shuldiner SR, Wong LY, Peterson TE, Wolfson J, Jermy S, Saad H, Lumbamba MAJ, Singh A, Shey M, Meintjes G, et al. Myocardial Fibrosis among Antiretroviral Therapy-Treated Persons with Human Immunodeficiency Virus in South Africa. Open Forum Infect. Dis. 2021;8.

32. Zanni M V., Awadalla M, Toribio M, Robinson J, Stone LA, Cagliero D, Rokicki A, Mulligan CP, Ho JE, Neilan AM, et al. Immune correlates of diffuse myocardial fibrosis and diastolic dysfunction among aging women with human immunodeficiency virus. Journal of Infectious Diseases. 2020;221:115–1320.

33. Tseng ZH, Moffatt E, Kim A, Vittinghoff E, Ursell P, Connolly A, Olgin JE, Wong JK, Hsue PY. Sudden Cardiac Death and Myocardial Fibrosis, Determined by Autopsy, in Persons with HIV. New England Journal of Medicine. 2021;384:2306–2316.

34. Peterson TE, Shey M, Masina N, Wong LY, Shuldiner SR, Wolfson J, Jermy S, Saad H, Lumbamba MAJ, Singh A, et al. Myocardial extracellular volume fraction is positively associated with activated monocyte subsets among cART-treated persons living with HIV in South Africa. Int. J. Cardiol. 2023;392.

35. Peterson TE, Hahn VS, Moaddel R, Zhu M, Fan J, De S, Haberlen SA, Palella FJ, Plankey M, Bader JS, et al. Human Immunodeficiency Virus–Associated Proteomic Signature of Myocardial Fibrosis and Incident Heart Failure. J. Infect. Dis. 2026;

36. Nelson MD, Szczepaniak LS, LaBounty TM, Szczepaniak E, Li D, Tighiouart M, Li Q, Dharmakumar R, Sannes G, Fan Z, et al. Cardiac steatosis and left ventricular dysfunction in HIV-infected patients treated with highly active antiretroviral therapy. JACC Cardiovasc. Imaging. 2014;7:1175–1177.

37. Sereti I, Krebs SJ, Phanuphak N, Fletcher JL, Slike B, Pinyakorn S, O’Connell RJ, Rupert A, Chomont N, Valcour V, et al. Persistent, Albeit reduced, chronic inflammation in persons starting antiretroviral therapy in acute HIV infection. Clinical Infectious Diseases. 2017;64:124–131.

38. Solages A, Vita JA, Thornton DJ, Murray J, Heeren T, Craven DE, Horsburgh CR. Endothelial Function in HIV-infected Persons. Clin. Infect. Dis. 2006;42:1325.

39. Bloomfield GS, Watanabe M, McCallum S, Aberg JA, Awwad A, Campbell TB, Cespedes MS, Chu SM, Currier JS, Diggs MR, et al. Heart Failure Risk and Events in People With HIV: The Randomized Trial to Prevent Vascular Events in HIV (REPRIEVE). Circ. Heart Fail. 2026;19.

40. Wise JM, Jackson EA, Kempf M-C, Oates GR, Wang Z, Overton ET, Siddiqui M, Woodward M, Rosenson RS, Muntner P. Sex differences in incident atherosclerotic cardiovascular disease events among women and men with HIV. AIDS. 2023;37:1661–1669.

41. Hanna DB, Ramaswamy C, Kaplan RC, Kizer JR, Daskalakis D, Anastos K, Braunstein SL. Sex- and poverty-specific patterns in cardiovascular disease mortality associated with human immunodeficiency virus, New York city, 2007-2017. Clinical Infectious Diseases. 2020;71:491–498.

42. Raghavan A, Rimmelin DE, Fitch K V., Zanni M V. Sex Differences in Select Non-communicable HIV-Associated Comorbidities: Exploring the Role of Systemic Immune Activation/Inflammation. Curr. HIV/AIDS Rep. 2017;14:220–228.

43. Ramirez MF, Pan AS, Parekh JK, Owunna N, Courchesne P, Larson MG, Levy D, Murabito JM, Ho JE, Lau ES. Sex Differences in Protein Biomarkers and Measures of Fat Distribution. Journal of the American Heart Association. 2024;13.

44. Savji N, Meijers WC, Bartz TM, Bhambhani V, Cushman M, Nayor M, Kizer JR, Sarma A, Blaha MJ, Gansevoort RT, et al. The Association of Obesity and Cardiometabolic Traits With Incident HFpEF and HFrEF. JACC Heart Fail. 2018;6:701–709.

45. Sax PE, Erlandson KM, Lake JE, McComsey GA, Orkin C, Esser S, Brown TT, Rockstroh JK, Wei X, Carter CC, et al. Weight gain following initiation of antiretroviral therapy: Risk factors in randomized comparative clinical trials. Clinical Infectious Diseases. 2020;71:1379–1389.

46. Shisana O, Labadarios D, Rehle T, Simbayi L, Zuma K, Dhansay A, Reddy P, Parker W, Hoosain E, Naidoo P, et al. South African National Health and Nutrition Examination Survey (SANHANES-1). Cape Town: HSRC Press. 2013

47. Beidelman ET, Rosenberg M, Wade AN, Crowther NJ, Kalbaugh CA. Prevalence of and Risk Factors for Peripheral Artery Disease in Rural South Africa: A Cross-Sectional Analysis of the HAALSI Cohort. J. Am. Heart Assoc. 2024;13.

48. Moyo-Chilufya M, Maluleke K, Kgarosi K, Muyoyeta M, Hongoro C, Musekiwa A. The burden of non-communicable diseases among people living with HIV in Sub-Saharan Africa: a systematic review and meta-analysis. EClinicalMedicine. 2023;65.

49. Yebyo HG, Günthard HF, Rehfuess EA, Serra N, Haile SR, Senn O, Lucas GM, Langselius O, Thorne JE, Marconi VC, et al. Statins for primary prevention of cardiovascular events in people with HIV: target trial and modelling study. BMJ Medicine. 2025;4:e001132.

50. Butt AA, Chang C-C, Kuller L, Matthew D, Goetz B, Leaf D, Rimland D, Gibert CL, Oursler KK, Rodriguez-Barradas MC, et al. Risk of Heart Failure With Human Immunodeficiency Virus in the Absence of Prior Diagnosis of Coronary Heart Disease. Arch Intern Med. 2011; 171(8): 737–743.

